# A district-level model review system to strengthen coverage and quality of Medical Certification of Cause of Death in India: Protocol for a population based feasibility and effectiveness study

**DOI:** 10.64898/2026.06.25.26356608

**Authors:** Madhusudan Muralidhar, Thilagavathi Ramamoorthy, Priyanka Das, Monesh B Vishwakarma, Sukanya Rangamani

## Abstract

**Background:** The current coverage of MCCD in India is only 22%. This is due to incomplete coverage of hospitals under MCCD and also lack of a system for non-institutional deaths in the country. The quality of MCCD in the country is also poor. One of the main reasons for this is the lack of review and feedback at the district level. This study would be the first of its kind study in the country to test the effectiveness and feasibility of involving the district level CRS/Health dept officials in review of MCCD

**Objectives:** To assess the feasibility and effectiveness of a district level review system for MCCD in improving the coverage and quality of MCCD

**Methods:** The study would be conducted in Chikkaballapura district for a period of 2 years. Local Registrars would do a first level of review of MCCD forms for completeness, use of abbreviations, legibility. They would also ensure that form 4/4A is written for all registered deaths in their area. A MCCD review committee would assess the quality of MCCD forms on a monthly basis and provide feedback to the certifying doctors. Comparison of the pre-test and post-test coverage and quality of MCCD will be done.

**Results:** Constitution of the audit committee, training of local registrars, doctors and committee members and baseline assessment have been completed. Intervention has been started from Nov 2025.

**Expected Outcomes:** Improved coverage and quality of MCCD and as a result cause of death data of the district

## Introduction

Medical Certification of Cause of Death (MCCD) is essential for accurate mortality surveillance and evidence-based decision-making. But the coverage of Medical Certification of Cause of Death (2022) in India out of the total registered deaths is 22% and there is a very slight increase in last decade.^1^ This is largely attributed to significant proportion of deaths (70%) occurring outside hospitals (deaths at home viz., deaths during sleep, sudden deaths etc., and those during travel to hospital) which are classified as non-institutional deaths. The cause of death in such cases is unlikely to be certified by any doctor because there is no medical attention received prior to death and/or there is no insistence on submission of cause of death certificate (form 4A) for death registration in case of non-institutional deaths. The Registration of Births and Deaths Act, 1969 and its amendment in 2023 states that for all deaths the ‘cause of death’ has to be certified by the doctor who attended to the decedent prior to his death, but does not clarify how cause of death can be certified in an individual who has not received any medical care in the recent past.^2^ In most of the states of India for non-institutional deaths, form 4A is neither insisted upon nor is there any suitable guidance provided. As a result of all these, non-institutional deaths (approximately 70% of all deaths) in India may be registered with the Local Registrar but may not have a medically certified cause of death. Also, out of all hospitals having inpatient facilities in our country 87% are covered under MCCD across different states but only 47.3% submit MCCD data to the Civil Registration System. As a result of all these, the MCCD coverage in India is dismal at 22%. ^1^

On the other hand, with regards to the quality of submitted MCCD forms in India, studies have shown that almost 100% of the forms have at least 1 error and >82% of the forms atleast one major error (which interferes with selection of underlying cause of death).^3-6^ Some of the interventions at the institutional and system level that have been attempted are training and use of software to facilitate capture of MCCD data in a standardized manner.^7,8^ However, a recent study conducted in 6 states of India by ICMR-NINE has revealed that the training conducted in states is infrequent and there is no review and feedback by CRS system/Dept of Health with regards to MCCD. Also, most of the respondents from the CRS had suggested that CRS system and Dept of Health at district level should be involved in the review of MCCD in order to improve the quality and coverage of MCCD (unpublished results).

In this context, the study aims to test the feasibility of having a district level review and feedback system for MCCD and its effectiveness in improving the coverage and quality of MCCD.

## Materials and Methods

### Study design and setting

This would be a ‘before and after comparison study’ to be carried out in Chikkaballapura district of Karnataka with a MCCD coverage of 12.2% which is below the Karnataka state average of 23.1%. Chikkaballapura is a district in the Southern Indian state of Karnataka with a population of 12.55 lacs.^9^ Mandya District of Karnataka with a similar MCCD coverage (11.8%) would be kept as control to compare the findings.^10,11^

The study shall be implemented by the PI, Co-PIs and the project staff from ICMR-NINE in collaboration with the Directorate of Economics and Statistics and Department of Health and Family Welfare, Karnataka as per the timelines in the table I and II. Preliminary meetings were held with Dept of Health and Family Welfare and Chief Registrar of Births and Deaths (Directorate of Economics and Statistics), Karnataka to obtain permission and their collaboration in this study. Subsequently, SOPs were developed and necessary directions to the Local Registrars, District Statistical Officers were issued from the Chief Registrar of Births and Deaths, Karnataka for reviewing and rectification of MCCD forms and to the PHC Medical Officers and other doctors of the district for certifying Cause of death for non-institutional deaths in their area by the Districtt Health Officer.

**Table I:** Timelines of baseline and endline assessment and interventions.

| Month 9 | Month 11 | Month 12 | Month 13 | Month 14 | Month 15 | Month 16 | Month 17 | Month 18 |
| --- | --- | --- | --- | --- | --- | --- | --- | --- |
| baseline assessment on month 8 data * | **Intervention period** |  |  |  |  |  |  | Post test assessment on month 17 data * |
|  | - | Assessment of month 11 data<sup>#</sup> | Assessment of month 12 data<sup>#</sup> | Assessment of month 13 data<sup>#</sup> | Assessment of month 14 data<sup>#</sup> | Assessment of month 15 data<sup>#</sup> | Assessment of month 16 data<sup>#</sup> |  |
|  | Review and rectification of MCCD by LR on ongoing basis |  |  |  |  |  |  |  |

<sup>#</sup>by the committee; \* by the investigating team

**Table II:**
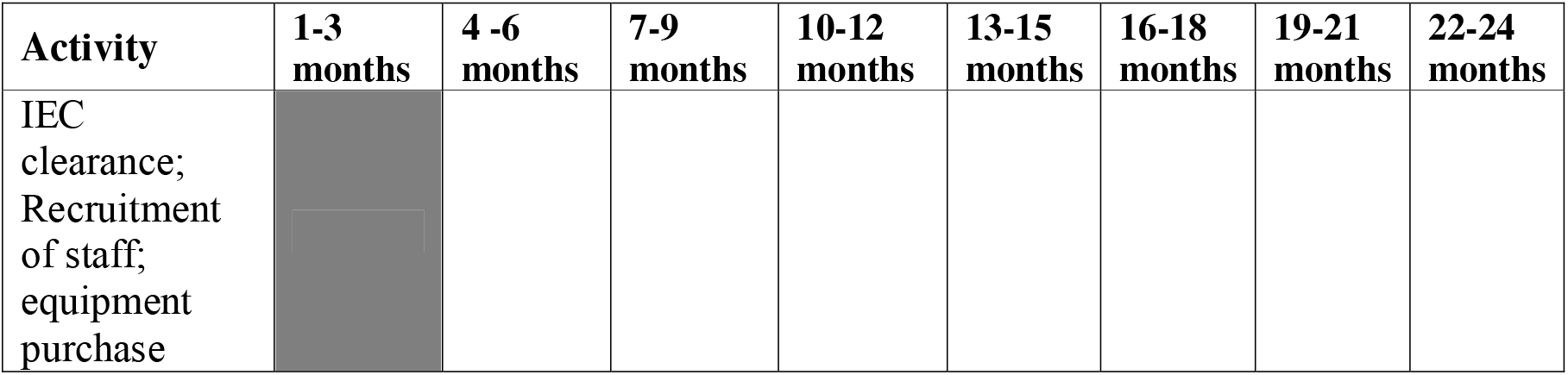
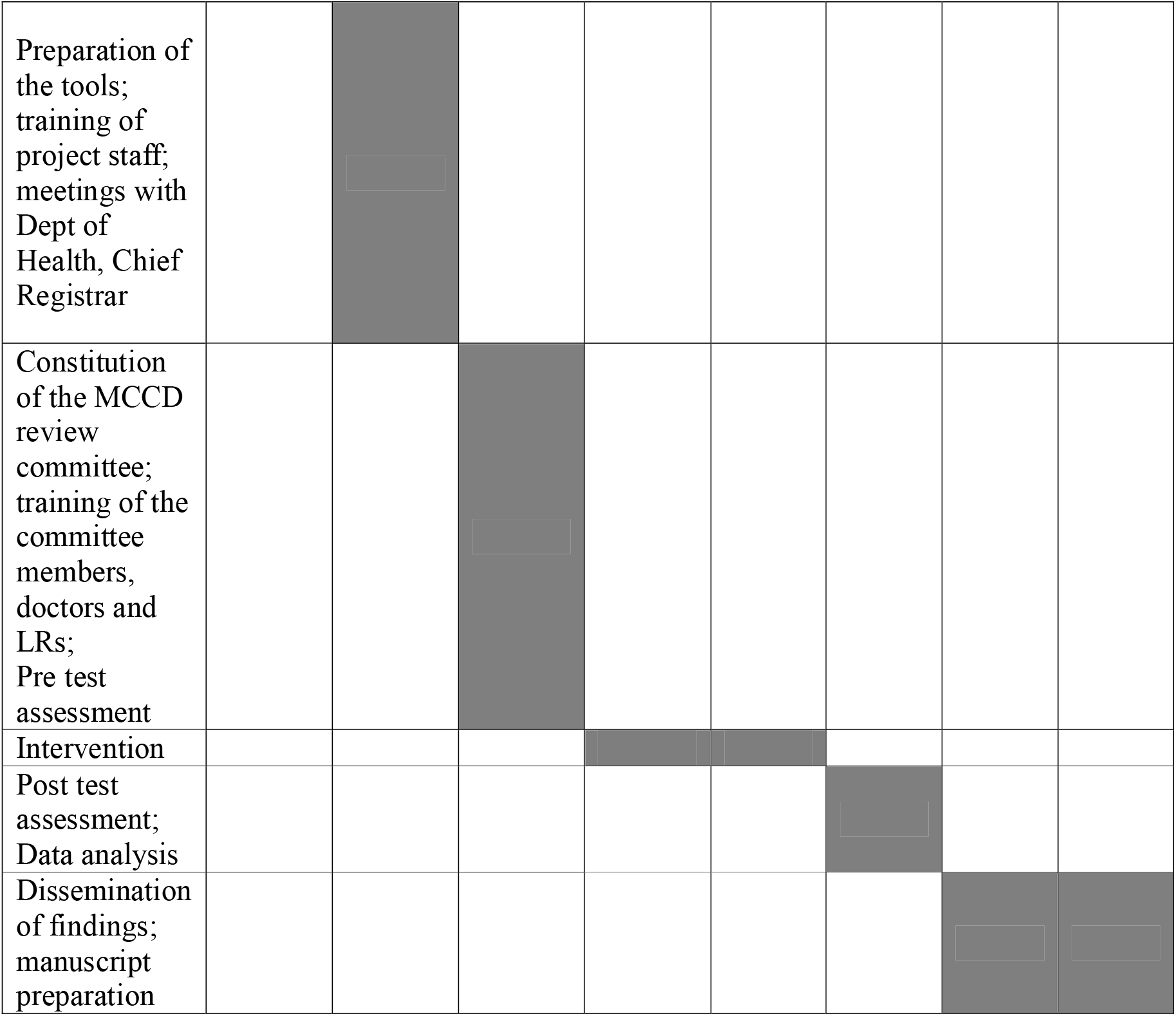
Timelines with achievable targets.

### Baseline assessment

Baseline assessment of the Quality and coverage of the MCCD were done prior to the start of intervention (table I). Assessment of quality of a sample of MCCD (obtained from the office of the district statistical officer) were done by MCCD e-audit tool which is the electronic version of the tool used in ‘Framework for audit of medical certification of cause of death at health facility’ using tabs.^12^ Assessment of Coverage of MCCD of the district was done based on the statistics available in the e-Janma portal (the CRS portal of Karnataka state).

### Interventions

The study will have 2 intervention components as follows:-

#### (a) Review by Local Registrars

An initial training was given to all Local Registrars (LR) of the district (LRs are the officials entrusted with the responsibility of registering deaths in their designated area under CRS. They include Panchayat Secretaries of Gram Panchayats, Village accountants of revenue circles, Health Officers/Heath Inspectors of urban local bodies and Resident/Administrative Medical Officers of govt hospitals) by ICMR-NINE to review the MCCD forms submitted to them for completeness of all fields, use of abbreviations in the Cause of Death section and legibility.^13^ The MCCD forms (both form 4 for institutional deaths and 4A for non-institutional deaths) submitted to the local registrar would be initially reviewed for completeness of all fields, use of abbreviations and legibility and in case of any mistakes returned to the certifying hospital/doctor for rectification and resubmission. The LR shall also ensure that all death report forms (form 2) submitted have form 4/4A. In case of non-institutional deaths where form 4A is not attached, the LR shall request the kin of the deceased to get form 4A filled by a doctor who has treated the deceased in his/her last illness (1st preference) or in the preceding 3 months and knows the medical history (2nd preference). In case the deceased has not received any such medical care in the preceding 3 months, then the LR shall request it to be got it done from the Administrative Medical Officer of the respective Primary Health Centre or Community Health Centre, Taluk Hospital or Resident Medical Officer of the District Hospital in this order of priority [who would be eliciting history using the Physician derived Cause of Death (PhyCoD) tool and arriving at cause of death]. Wherever MCCD could not be done the LR would record the reason for the same. All the AMOs of the PHC, CHC, Taluk Hospitals and RMOs of the district have been initially trained in MCCD and deducing of cause of death in non-institutional deaths using the PhyCoD tool.^14^ Instructions have been issued by the District Health Officer (DHO) to all the doctors of the district for co-operation in this regard. In this manner, the LR would ensure MCCD is done for all the deaths registered by him/her. A Key Informant Interview of a sample of LRs and the PHC, CHC, Taluk Hospital AMOs, RMOs who have certified Cause of Death for non-institutional deaths would also be done in the end to understand the various challenges and enablers faced by them during the intervention.

#### (b) Review and feedback by a committee

A Committee of doctors from the 2 Medical Colleges of the district (1 government and 1 private) has been constituted by the District Health Officer. The committee consists of clinicians from major clinical departments (like General Medicine, General Surgery, Obstetrics and Gynecology, Pediatrics, Emergency Medicine etc.,), Forensic Medicine, Community Medicne and the district statistical officer (officer in-charge for CRS/MCCD in the district). The committee members were trained in MCCD and International Statistical Classification of Diseases and Related Health Problems (ICD) and also oriented about the review process. This committee would meet at monthly intervals and review a sample of the forms of the previous month and give feedback to the certifying doctors/hospitals about the errors observed. The committee would also review the cause of death data of other portals like Integrated Disease Surveillance Programme (IDSP), Integrated Health Information Platform (IHIP), NIKSHAY portal (for Tuberculosis deaths), RCH portal (for maternal, infant and neonatal deaths) etc., and check for consistency and also coverage of MCCD of the district from the e-janma portal.

The PI and other investigators will liaise between district health, CRS officials, audit committee and LRs and doctors in the periphery for implementation of the interventions, providing inputs for any mid-course corrective measures following the monthly audits.

The above 2 interventions would be carried out for 6 months.

Endline assessment of the Quality and coverage of the MCCD would be done on the month succeeding the end of intervention on the previous month data (table 1).

### Outcome measures

The indicators for measuring the quality of MCCD would be proportion of errors of each type (minor: viz., incompleteness of various fields, use of abbreviations in the cause of death section etc., major: wrong sequence of events, vague/ill-defined terms in the cause of death section etc.,) and also the aggregate quality score. The aggregate score takes into account the number of errors of each type in each form assessed.^11^MCCD coverage would be calculated based on the proportion of deaths for which MCCD was done out of total registered deaths which would be done based on the statistics available in the e-Janma portal.

Other secondary indicators for measuring the outcome of the intervention would be:-

a. Change in the knowledge/skill level of the LRs, PHC MOs and district level review committee members assessed by pretest (before training) and post test (post training).
b. Proportion of LRs and MOs who participated in the training
c. Proportion of deaths for which review was done by LRs out of total registered deaths
d. Proportion of form 2s initially without MCCD which were sent back and certified subsequently by the doctor out of those which didn’t have MCCD initially
e. Proportion of form 4/4As for which the LR attempted to rectify the errors

#### Sample size

For the baseline, endline and monthly assessments during the intervention period, 5% of the forms of the preceding month selected by systematic random sampling would be considered. Chikkaballapura district had registered 10,009 deaths in 2023, which would be approximately 834 deaths/month. Hence, the committee would assess 5% of 834 i.e., 42 or 50 forms whichever is more, every month. Chikkaballapura district has 1583 registration units. Assuming 5% vacancies of LRs, each LR would be required to assess approximately 3 forms during the 6 months period.^15^

### Tools for data capturing and management

#### (i) Local Registrar

A web-based tool to capture data at the Local Registrar level has been developed by ICMR-NINE. Access will be provided to LRs through valid credentials provided by ICMR-NINE. The tool would contain fields to capture basic details like name of the registration unit, district, place of death etc., followed by a check list to look for various errors. In case of non-availability of from 4/4A initially, the portal has the facility to save the record in draft mode, so that it can be retrieved subsequently once the form 4/4A becomes available. After completion of the review, the errors in the form 4/4A will be highlighted and option would be provided to LR to save the record in draft mode to provide an opportunity to rectify the errors. In case of non-availability of MCCD form or inability to rectify the errors even after due efforts, option has been provided to record the reasons for the same and submit the record, without interference with the death registration process.

Option to search records has also been provided. Data will be saved in the ICMR-NINE cloud. The tool will be used by the Local registrar in his/her system (desktop).

#### (ii) Audit Committee

A web based responsive tool would be developed for the audit committee members to review the MCCD forms and record the various clerical, certification and coding errors based on a checklist. Access will be through valid credentials provided by ICMR-NINE. The tool will also provide the facility to generate the error-wise report and aggregate quality score hospital-wise, registration unit-wise, taluk-wise and and also for entire district using which we can track the progress.

Data will be encrypted using standard encryption algorithm. A dashboard will be developed to monitor the progress of the work and data transmission. Regular syncing of data with the local ICMR-NINE server will be done. The technologies that would be employed are ASP.NET, VB.NET, HTML, CSS, Bootstrap 4.0 and database used would be Microsoft SQL Server 2016.

##### Statistical analysis

Data will be analysed using appropriate statistical software. MCCD coverage will be calculated as the proportion of deaths with medical certification out of total registered deaths using data from the e-Janma portal. Pre- and post-intervention coverage will be compared using the Chi-square test, and absolute change will be reported.

For quality assessment, the proportion of forms with each type of minor and major error will be calculated, along with the mean aggregate quality score. Differences in proportions of error types between baseline and endline will be assessed using the Chi-square test, and differences in mean aggregate scores will be compared using independent t-test or non-parametric equivalents, as appropriate.

Pre- and post-training knowledge scores will be compared using paired t-test or Wilcoxon signed-rank test. Secondary process indicators will be expressed as proportions and compared using the Chi-square test. A p-value <0.05 will be considered statistically significant, and all tests will be two-sided.

##### Ethical considerations

Ethical clearance from the ICMR-NINE Institutional Ethics Committee has been obtained (No: NCDIR/IEC/3084/2024 dated 30/01/2025). Identification details of the deceased will not be captured. Data would be stored in password protected systems and would have access only by PI/Co-PIs. The review details/feedback on MCCD would be shared with the respective hospitals/doctors only and not with other hospitals.

##### Peer Review and approvals

The proposal has been initially peer reviewed by 2 experts working in the field and subsequently approved by Scientific Advisory Committee of ICMR-NINE (PI’s institute) and Project Review Committee of the ICMR (Funder).

## Results

As on the date of submission (i.e., 17/02/2026), the following have been completed (fig 1):-

**Fig 1:**
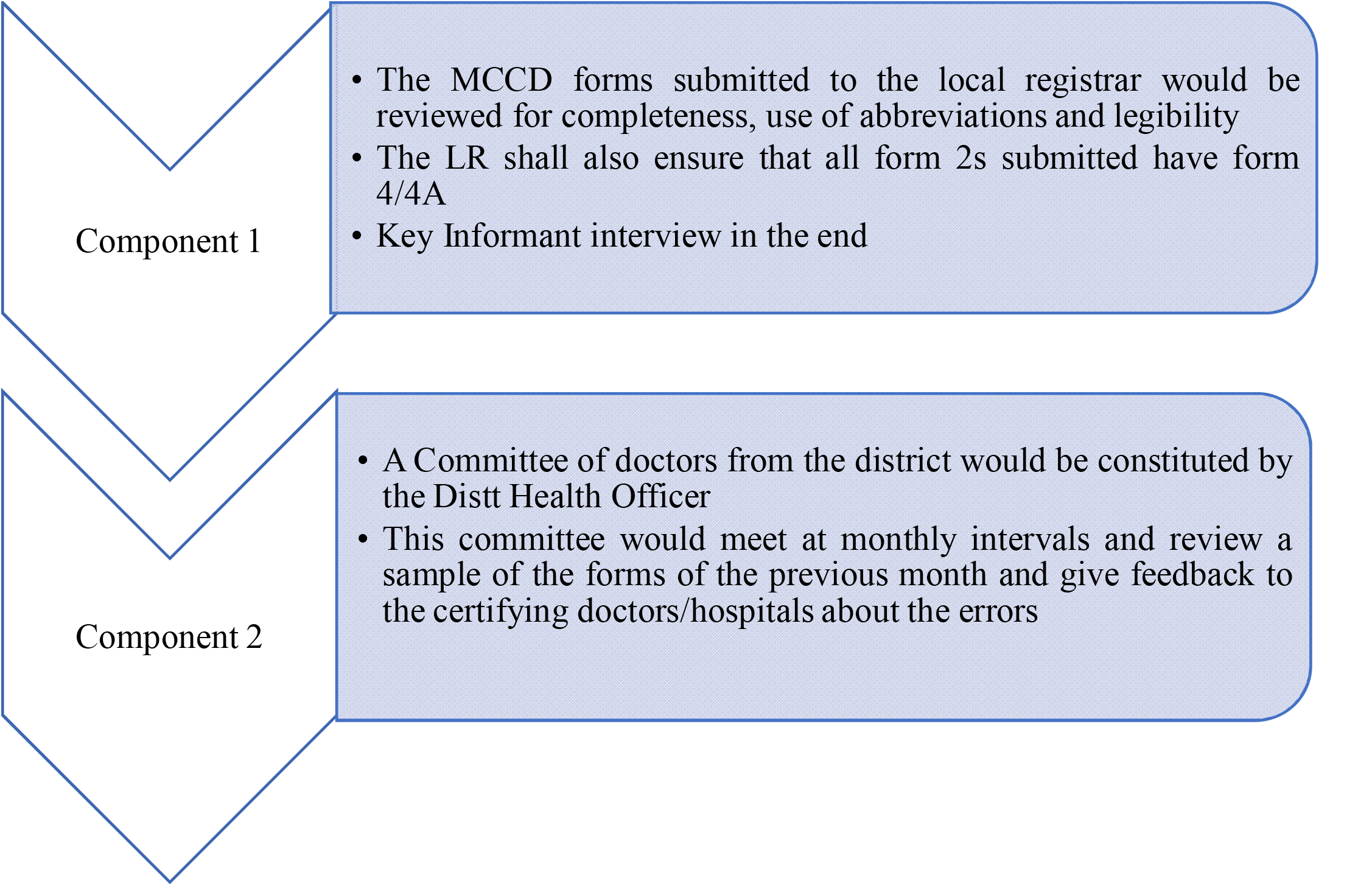
Methodology of the study.

a. IEC clearance (Jan 2025)
b. Equipment purchase (Feb-Mar 2025)
c. Recruitment of staff (Jan-Apr 2025)
d. Training of project staff (May 2025)
e. Preparation of the tools (Apr-Aug 2025)
f. Meetings with Dept of Health, Chief Registrar, Directorate of Medical Education and identification of nodal officers (Mar-Jun 2025)
g. Meetings with District Health Officer, District Statistical Officer (Jun 2025)
h. Constitution of the District MCCD review committee (Sep 2025)
i. Training of the committee members, doctors and LRs (Sep 2025)
j. Baseline assessment in Chikkaballapura and Mandya (Sep 2025)
k. Start of intervention (Nov 2025 onwards)

308 LRs, 41 doctors (Administrative Medical Officers of PHC, CHC and Taluk hospitals and Resident Medical Officer of the district hospital) of the district and 19 members of the audit committee were trained during the training. 3 audit committee meetings have been held till date and the minutes of the meeting have been shared with stakeholders for action.

## Discussion

MCCD is not only a technical necessity but a high-return on investment with respect to national development. Strengthening MCCD within CRVS systems is foundational for achieving Sustainable Development Goals and building resilient, data-driven governance.^16^ This study intends to address the gap of coverage as well as quality of MCCD in the country by developing a system for review and feedback by involving the *existing functionaries* in the district which is likely to be feasible and sustainable. All the deaths in a district would be registered through Local registrars. Since local registrars are routinely involved in registering deaths and cause of death information in the CRS portal involving them in reviewing whether all form 2s have form 4/4As and also the completeness, use of abbreviations and legibility is likely to be feasible, efficient and effective.

Although attempts have been made to cover non-institutional deaths by Verbal Autopsy under Sample Registration System (SRS), the limitation of VA is that history is collected by a lay person (SRS supervisor) and subsequently reviewed by a medical doctor (who is located far away and is not aware of the context) to arrive at cause of death. Because of these, there are chances of information being lost or misinterpreted. Also, the history collected in VA under SRS is in the local language. Hence, a considerable number of medical doctors proficient in various regional languages need to be available. This is a challenge in the case of some regional languages. In addition, because of the complicated process, there is a considerable delay in arriving at cause of death (approximately 9 months).^17,18^ The current study attempts to address these gaps by involving the local doctors from within the system in MCCD. Hence the limitations of language, delay, and miscommunication are likely to be minimised. Further since medical doctors are routinely involved in the history taking and diagnosis, the cause of death arrived at compared to VA is likely to be more accurate. It is universally accepted that MCCD is the gold standard for arriving at cause of death and verbal autopsy is only an interim arrangement till CRVS systems of countries are developed enough to cover all deaths under MCCD and covering all deaths under MCCD shall be the ultimate aim of all countries.^19^ In this context, the study attempts to develop a system to improve the coverage and quality of MCCD. Studies have shown that improving certification quality can lead to more actionable mortality data.^16^

However, a few challenges have been encountered during the initial phase of implementation like:-

- Resistance by the doctors to issue MCCD in non-institutional deaths because of the potential medico-legal issues. In discussion with DHO and audit committee, it has been decided to print on the form 4As the following phrase:-“the cause of death provided is provisional and based on the *available* information and is for Civil Registration System purposes only. The final cause of death can be obtained only after an autopsy”. The doctors have also been instructed to write the phrases such as ‘alleged history of’, probable etc., wherever they are not sure of the cause of death in order to allay their apprehension.
- Resistance from the kin of the deceased to get MCCD done, because of the need to travel to get MCCD and also their limited understanding about the importance of MCCD
- Difficulty to reach to all the local registrars of the district as they are from different departments like Revenue, Rural Development and Panchayat Raj, Health etc.,

The findings of the study including the challenges faced shall be shared with the Chief registrar of Births and Deaths, Karnataka, Dept of Health, Karnataka and Office of the Registrar General of India (ORGI). Based on all their inputs, the approach would be suitably modified and advocated for implementation in other districts and states of the country to increase the coverage and quality of MCCD.

## Data Availability

All data produced in the present study are available upon reasonable request to the authors

## Acknowledgements

The authors would sincerely acknowledge the guidance and support provided by the Director, ICMR-NINE, Bengaluru, administrative support provided by the Administrative staff of ICMR-NINE, co-operation and support of the Dept of Health and Directorate of Economics and Statistics, Karnataka.

## Data Availability

The data will be deposited into the institutional data repository of the Indian Council of Medical Research-National Centre for Disease Informatics and Research (ICMR-NINE), and access to other researchers will be provided subject to approval of the competent authority.

## Authors’ Contributions

Conceptualization: MM, SR,

Funding acquisition: MM

Methodology: MM, SR

Software: PD, MBV

Writing – original draft: MM, SR, TR

Writing – review & editing: MM, SR, TR

